# Early warning signals observed in motor activity preceding mood state change in bipolar disorder

**DOI:** 10.1101/2024.03.14.24304269

**Authors:** Petter Jakobsen, Ulysse Côté-Allard, Michael Alexander Riegler, Lena Antonsen Stabell, Andrea Stautland, Tine Nordgreen, Jim Torresen, Ole Bernt Fasmer, Ketil Joachim Oedegaard

**Affiliations:** Division of Psychiatry, Haukeland University Hospital, Bergen, Norway; Department of Clinical Medicine, University of Bergen, Bergen, Norway; Department of Technology Systems, University of Oslo, Oslo, Norway; SimulaMet, Oslo, Norway; Department of Global Public Health and Primary Care, University of Bergen, Bergen, Norway; Department of Informatics, University of Oslo, Oslo, Norway

**Keywords:** Bipolar Disorder, Mood Disorders, Recurrence, Motor Activity, Nonlinear Dynamics, Systems Analysis, Unsupervised Machine Learning

## Abstract

**Background:** Alterations in motor activity are well-established symptoms of bipolar disorder, and time series of motor activity can be considered complex dynamical systems. In such systems, early warning signals (EWS) occur in a critical transition period preceding a sudden shift (tipping point) in the system. EWS are statistical observations occurring due to a system’s declining ability to maintain homeostasis when approaching a tipping point. The aim was to identify critical transition periods preceding bipolar mood state changes.

**Methods:** Participants with a validated bipolar diagnosis were included to a one-year follow-up study, with repeated assessments of the participants’ mood. Motor activity was recorded continuously by a wrist-worn actigraph. Participants assessed to have relapsed during follow-up were analyzed. Recognized EWS features were extracted from the motor activity data and analyzed by an unsupervised change point detection algorithm, capable of processing multi-dimensional data and developed to identify when the statistical property of a time series changes.

**Results:** Of 49 participants, four depressive and four hypomanic/manic relapses among six individuals occurred, recording actigraphy for 23.8 ± 0.2 hours/day, for 39.8 ± 4.6 days. The algorithm detected change points in the time series and identified critical transition periods spanning 13.5 ± 7.2 days. For depressions 11.4 ± 1.8, and hypomania/mania 15.6 ± 10.2 days.

**Conclusion:** The change point detection algorithm seems capable of recognizing impending mood episodes in continuous flowing data streams. Hence, we present an innovative method for forecasting approaching relapses to improve the clinical management of bipolar disorder.

## Background

Bipolar disorder is a severe and incapacitating mental disorder of unknown etiology, characterized by recurrent and alternating mood episodes of mania and depression.^1^ The main goal of treatment is to avoid future mood episodes, through measures such as mood-stabilizing medications, lifestyle modifications, and alertness to early signs of new impending episodes.^2^ Although such early signs are usually consistent with subthreshold symptoms and observable for several weeks, they are habitually neglected.^3^ Consequently, developing a reliable method for forecasting an impending mood episode has the potential to revolutionize the management of bipolar disorder.^4^

Alterations in motor activity are essential symptoms of bipolar disorder.^5^ Time series of motor activity contain observations of mood and energy fluctuations, and the complex recurring circadian interplay of internal biological clocks and external social rhythms.^6,7^ In complex dynamical systems, early warning signals (EWS) occur in a critical transition period preceding a sudden noticeable shift in the system, often referred to as a tipping point.^8–10^ EWS are statistical time series observations that occur when a dynamic system’s ability to maintain homeostatic equilibrium begins to diminish as it approaches a tipping point.^11^ Hypothetically, this model could have universal applicability, potentially linking a diverse range of phenomena. These include geological shifts such as those in earthquakes, ecological dynamics within ecosystems, hydrodynamics in ocean circulation, meteorological patterns in climate, and pathophysiological shifts in chronic human diseases — notably, epileptic seizures, acute asthma attacks, and cardiovascular events.^12–14^

Within the field of mental health, it has been hypothesized that adopting a dynamical system perspective holds great innovative potential for the overall understanding, treatment, and management of mental disorders.^15^ Evidence supports this hypothetical model; specifically, EWS indicators such as increased variance and autocorrelation in self-rated mood have been observed preceding shifts from stable to unipolar depressed mood.^16^ Furthermore, a psychotherapeutic study observed EWS preceding changes in clinical mood in daily self-ratings from affective patients, by using a mathematical dynamic complexity algorithm.^17^ Regarding bipolar disorder, a study of patients experiencing mood transitions investigated the manifestation of critical transitions in daily self-reports.^18^ Their findings were optimistic, as alterations in autocorrelation and variance forecasted upcoming relapses. However, this study also found that the absence of EWS did not necessarily predict the absence of a tipping point.

Finally, a study utilizing artificial intelligence to analyze social media postings was able to predict clinically validated depression three months in advance, albeit with only moderate accuracy.^19^ A unifying aspect of all these studies is that they were based on data assembled from either participant or clinician-rated assessments. Such subjective data may be biased by personal opinions, understandings, or interpretations. In contrast, objective data sources, such as motor activity, contain more accurate and unbiased information.^5^ Promisingly, a single-case motor activity study spotted augmented variance in advance of a switch from depression to hypomania.^20^

Additionally, a systematic review of dynamical system models in bipolar disorder included 26 studies of various methodologies.^21^ While most focused on mood dynamics, some studies offered mathematical simulations connecting disturbed circadian rhythms to relapses in bipolar disorder. Overall, the papers presented hypothetical models lacking empirical support, aside from nine studies that analyzed subjective data on mood, sleep, and perceived energy, looking for patterns in nonlinear mathematical models of mood, rhythmicity, and oscillation frequencies. The reviews’ overall conclusion: evidence-based longitudinal studies on motoric and digital biomarkers are in demand.

Disturbed circadian rhythmicity has been proposed as the driving force behind the transition to mood state relapse.^22^ The association between disrupted circadian rhythmicity and bipolar mood episodes is well-established,^7^ entrenched by the strong association between sleep disturbance and mood episodes.^23^ However, associations do not necessarily imply causality. Although the sleep-wake cycle is mainly under circadian control, and closely connected to the suprachiasmatic nucleus (SCN) controlled rest-activity and light-dark cycles,^6^ this arrangement mainly regulates the body’s need for sleep. However, the probability of falling asleep, and the quality of sleep appear to be controlled by other non-circadian factors.^23^ In this regard, human consciousness is proposed as a substantial non-circadian factor controlling sleep.^24^

The human brain is not fully understood, but mood, motivation, and energy are considered outcomes of internal or environmental inputs processed and assessed in the brain.^25^ A central component is the limbic system - an interconnection of deep brain structures, involved in instituting the emotional state, recalling learned experiences, storing new memories, and establishing levels of anxiety and motivation. It incorporates the brain circuits controlling reward and motivation,^26^ a system associated with bipolar mood.^27^ Accordingly, the limbic process establishes the emotional and motivational state of the consciousness, corresponding to the central nervous system’s (CNS) level of arousal.^28^ Immediate CNS arousal related to fear and strong feelings have been observed in motor activity. A technological study of accelerometer-equipped soccer supporters found significant differences between periods containing goals to non-event periods in the match, analyzing high resolution uncompressed actigraphy data.^29^ Likewise, causality between fear and behavioral pattern changes have been observed in studies of laboratory mice.^28^ This evidence supports the hypothesis proposed in a systematic review on activation in bipolar disorder;^5^ namely, that motor activity can be considered an objective observation of the inner psychological state expressed in behavior patterns.

Communication from the CNS to the rest of the body occurs via two branches of the peripheral nervous system. The first, the autonomic nervous system, controls involuntary actions and can be observed through digital biomarkers. The second, the somatic nervous system, governs voluntary muscle movement and is observable in motor activity. A central regulator between the two is the hypothalamus; a small nucleus in the limbic system that, besides hosting the SCN, is considered the communication center of the CNS.^30^ Its primary objective is to maintain stability and safety in bodily functions and rhythms, ensuring they remain at a homeostatic equilibrium. Our research group has previously observed relationships between autonomic and somatic signals, when comparing hospitalized manic patients to euthymic humans with bipolar disorder, in an advanced machine learning analysis of multisensory data.^31^ Furthermore, when we compared a selection of hospitalized manic patients to their remitted selves, we found reduced heart rate variability in mania, indicating autonomic dysregulation,^32^ in addition to increased complexity and reduced variance in manic motor activity patterns.^33^

Bipolar disorder can be conceptualized as three distinct states, where euthymic individuals are episodically driven towards either depression or hypomania/mania ((hypo)mania). Drawing from this basic model and acknowledging the primary role of the hypothalamus in maintaining homeostasis,^30^ we propose that its function serves as a continuous signal, fluctuating around the euthymic equilibrium. As such, when the system approaches a tipping point, EWS should be observable, persisting for a duration before a rapid shift stabilizes the system around a new equilibrium of either depression or (hypo)mania.

Change point detection (CPD) algorithms are developed to identify moments where the statistical property of a time series changes.^34^ However, developing effective CPD algorithms for forecasting impending mood episodes in motor activity is ambitious due to the complexity of analyzing prolonged and multimodal time series data sampled at a high frequency. Consequently, a scalable algorithm that can handle large data volumes and integrate multiple data features into a cohesive analysis is necessary. Moreover, such an algorithm must be capable of operating in near real-time to provide timely insights for interventions and treatment, crucial for successful clinical implementation of the model. The Latent Space Unsupervised Semantic Segmentation (LS-USS) algorithm is a state-of-the-art scalable unsupervised domain agnostic change point detection algorithm capable of processing multi-dimensional time series,^35^ and has a presumed potential for demonstrating the feasibility of using objective, non-invasive biomarkers in the form of motor activity data for forecasting mood state transitions in bipolar disorder.

Assuming that an equilibrium signal from the hypothalamus is observable in time series of motor activity, we hypothesize that the application of the LS-USS algorithm to motor activity data will reveal distinct patterns of change, characterized by alterations in statistical EWS indicators, prior to the onset of mood state changes. Accordingly, this study aimed to identify critical transition periods preceding bipolar mood state changes.

## Patients and methods

### Participants

Eligible patients were identified from a pool of participants in the outpatient part of the two-phased detecting and predicting mood transitions in bipolar disorder study at Haukeland University Hospital, Bergen, Norway.^33^ The first phase of the study included patients admitted to Haukeland University Hospital with agitated features of an ongoing manic or depressed episode. The second phase was designed as a one-year outpatient follow-up study. Participants were either continued from the first phase when discharged, enrolled from the hospital’s outpatient clinic, or recruited from a local advocacy group for bipolar disorder.

During the one-year follow-up nine regular clinical follow-up meetings, where the participants’ mood state was assessed by clinical interviews took place. Participants who were assessed to be in a new mood episode at a follow-up visit, meeting the following criteria, were included in the final analysis: 1) The mood change must have appeared between two clinical assessments no more than 60 days apart, and 2) The participant was euthymic at the first assessment point, and in an ongoing mood episode at the next clinical meeting point.

Inclusion criteria for the study were Norwegian-speaking individuals between 18 and 70 years diagnosed with a bipolar disorder diagnosis according to ICD-10, able to comply with instructions, and with an IQ clinically evaluated to be above 70. Exclusion criteria were previous head trauma needing hospital treatment, having an organic brain disorder, substance dependence (excluding nicotine), or being in a withdrawal state. Participants were included in the present study between September 25th, 2018, and July 6th, 2020. The study protocol was approved by The Norwegian Regional Medical Research Ethics Committee West (2017/937). Informed, written consent was obtained from all participants, and no compensation for participation was given.

### Clinical Assessments

Patients’ mood states were evaluated at all assessment points using the Young Mania Rating Scale (YMRS)^36^ and the Montgomery Asberg Depression Rating Scale (MADRS)^37^. The threshold for euthymia was defined as a sum score of nine or below for YMRS,^33^ and below 12 for MADRS, a threshold commonly utilized by randomized controlled trials of bipolar depression.^38^ Diagnoses were validated at the baseline visit by the Norwegian version of the Mini International Neuropsychiatric Interview (M.I.N.I.) version 6.0.0.^39^ Furthermore, all relapsed participants were comprehensively questioned about their view on when (specific date or period) their change in mood had occurred.

### Recording and processing of the motor activity time series

Motor activity was recorded with the wrist-worn GENEActiv actigraph (Activinsights, Cambridge, United Kingdom), containing a 3-axis accelerometer module that measures acceleration in gravitational force equivalents (g).^40^ The accelerometers were set to record at a 10 Hz sampling frequency, to minimize the load on the battery and increase the recording length to 60 days. We told the participants to wear the actigraph on the wrist of their choice continuously, i.e., for 24 hours a day/7 day a week. At the clinical follow-up meetings, the accelerometers were charged, and the raw data files extracted using the GENEActiv PC Software (ver. 3.2).

The raw data files were processed in the statistical software RStudio version 1.2.1335. For each time series of motor activity, the absolute sum of vector magnitudes was calculated for the 3-axis’ activity counts per Hz, by the formula |SQRT (x^2^ + y^2^ + z^2^) – Gravity|, and the data was stored in one-second epochs. All data recorded before the first, and after the last complete 24-hour cycle (midnight to midnight) were ignored, to only access complete 24h cycles in our time series analyses. Furthermore, to evaluate the quality of the collected time series, and to detect potentially biasing non-wear periods, we explored the raw unprocessed GENEActiv files with the Research Community-Driven open source R-package GGIR (ver. 2.7-1).^41^ Importantly, GGIR was utilized solely for quality assessment of the data. No data was imputed into the time series, and none of the output estimates on activity and sleep from the package were included in or utilized by the CPD algorithm.

To finalize the preparation of the motor activity data for the CPD algorithm, the one-second epochs were segmented into non-overlapping intervals of 300 samples, corresponding to five minutes per segment. For each non-overlapping window, the estimates of mean activity, coefficient of variation (CV), root mean square of successive differences (RMSSD), autocorrelation at lag 1, skewness, and sample entropy were computed. Skewness expresses the degree of asymmetric distribution in time series. Autocorrelation evaluates pattern repetitiveness, and at lag 1 determines the correlation of a time series with itself delayed one interval. The two estimates of variance, CV and RMSSD, articulate the stability of a time series’ mean. Sample entropy measures the complexity, randomness, and predictability of patterns.^5,33^ All features except mean activity are recognized EWS indicators.^9,11^ Consequently, the actigraphy signal from each participant was represented by a 6xY matrix, where Y represented the number of five-minute epochs in the time series.

### The Latent Space Unsupervised Semantic Segmentation algorithm

The Latent Space Unsupervised Semantic Segmentation (LS-USS) algorithm is a domain-agnostic and scalable CPD algorithm that can be applied to both online and offline datasets.^35^ LS-USS is based on the Fast Low-cost Online Semantic Segmentation (FLOSS) algorithm.^42^ The central hypothesis behind the development of the LS-USS algorithm is that similar segments are more alike than subsequences occurring after a change point. The algorithm works by learning a mapping between each subsequence in the time series to the most similar, non-trivial (i.e., sufficiently far away so that it cannot match with itself or part of itself) subsequence in the time series. The mapping between two subsequences is represented as a directed arc linking the centroid point of subsequence A with the centroid point of subsequence B. Hence, CPD can be performed by identifying the point in the time series where the minimum number of arc crossings occur over a particular point.

The LS-USS algorithm is composed of three main components; an autoencoder, applied to map the multi-dimensional subsequences into a lower-dimensional space, The Latent Space Matrix Profile, linking the non-trivial subsequences together in a computationally efficient manner, and the Corrected Arc Curve (CAC), used for identifying change points. Note that the term “Corrected” is used to refer to the fact that the construction artifacts resulting from the use of windows have been accounted for, to avoid detecting false change points on the border of the CAC. Change points are identified by recognizing the local minimum value data point in the CAC, and by using an exclusion zone, corresponding to a relevant and reasonable amount of time, to avoid detecting change points inappropriately close to each other. We selected an exclusion zone of 3-day as a conservative estimate. This choice was based on observations of EWS more than 15 days preceding the onset of hypomania in the only prior study of bipolar mood relapse analyzing motor activity.^20^ Similar timeframes were presented in the study observing EWS in daily self-reported data.^18^ For the sake of concision, we defer the description of the Latent Space Matrix Profile and the CAC components of LS-USS to the original paper.^35^

In this study, we introduced a compact Convolutional InceptionTime Network-Long Short-Term Memory (CIN-LSTM) architecture as the autoencoder component of the LS-USS algorithm, designed to distill the information present within the multi-dimensional motor activity time series.^35^ During training, the network is conditioned to preserve the most crucial information for reconstructing the time series, ensuring it captures a meaningful representation of the multi-modal time series within a diminished dimensionality. The architecture employs a Convolutional InceptionTime Network (CIN) to discern features across varying time scales in the data, while the LSTM component, receiving these features from the CIN, focuses on identifying sequential information across them. Ultimately, this architecture facilitates the learning of a 1D mapping from a multi-modal time series of relevant features. The autoencoder design is presented in Figure 1. The CIN-LSTM encoder module contains two inception blocks followed by the LSTM network. Similarly, the decoder modules contain first an LSTM network followed by two Inception blocks. RangerLars^43^ is employed for the network optimization with batch size of 128. To prevent potential data leakage, we utilized a random search with 50 candidates (each evaluated five times) to select the optimal learning rate (lr=0.0037). This hyperparameter search was performed on actigraphy data from bipolar disorder patients not included in this study to avoid potential data leakage. The random search was performed using a logarithmic scale between 10∧-6 and 1, following a uniform distribution. To train the autoencoder, we considered a window size of 144 samples, each sample representing a 5-minute interval, corresponding to a 12-hour duration. The learning rate was annealed using the 1 cycle policy, as described by Smith & Topin,^44^ over a total of 500 epochs. During the training phase, the performance of the autoencoder was assessed using the Mean Squared Error (MSE) loss function. The MSE provides a quantitative measure of the differences between the original input sequence and the autoencoder’s reconstructed output. During training, the weights of the network are iteratively adjusted to minimize the MSE loss across the training dataset.

**Fig. 1.**
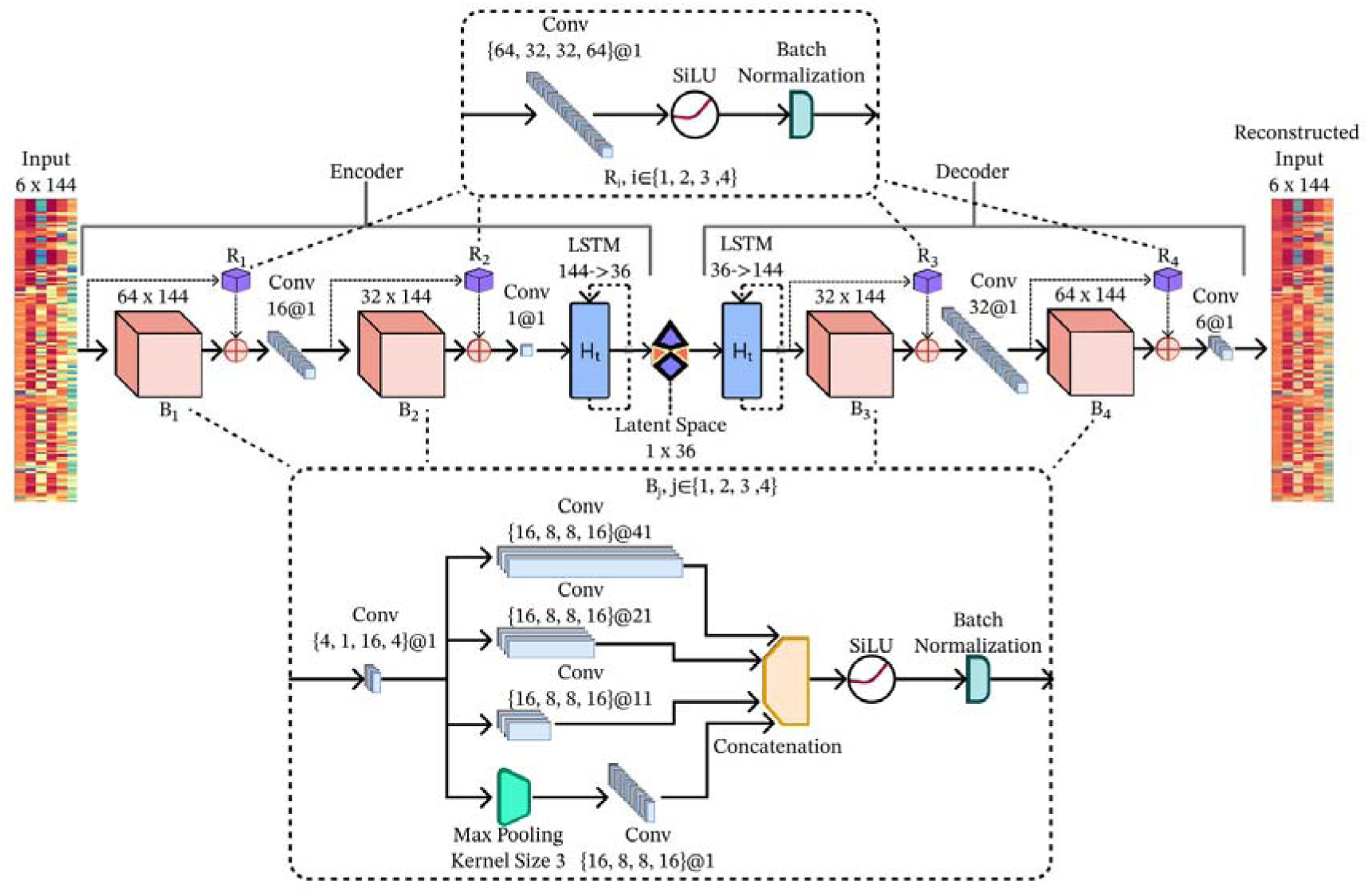
The autoencoder CIN-LSTM architecture employed for change point detection. The architecture contains 156 968 learnable parameters. R_i_ refers to the ith residual block (I ∈ {1, 2, 3, 4}, while B_j_ refers to the Jth InceptionTime block (j ∈ {1, 2, 3, 4}). Conv refers to a convolutional layer, while LSTM to a Long short-term memory network. Finally, the plus signs refer to an element-wise summation.

### Statistics

The statistical software RStudio version 2023.06.2 was used for designing Fig. 2, which presents the Corrected Arc Curves, the detected change points, as well as the self-reported possible occurrences of mood changes. SPSS version 29.0.0.0 was used to calculate the results presented in Tables 1 and 2, and to perform Paired Samples significance testing (two-tailed), considering p<0.05 statistically significant.

## Results

**Table 1.**
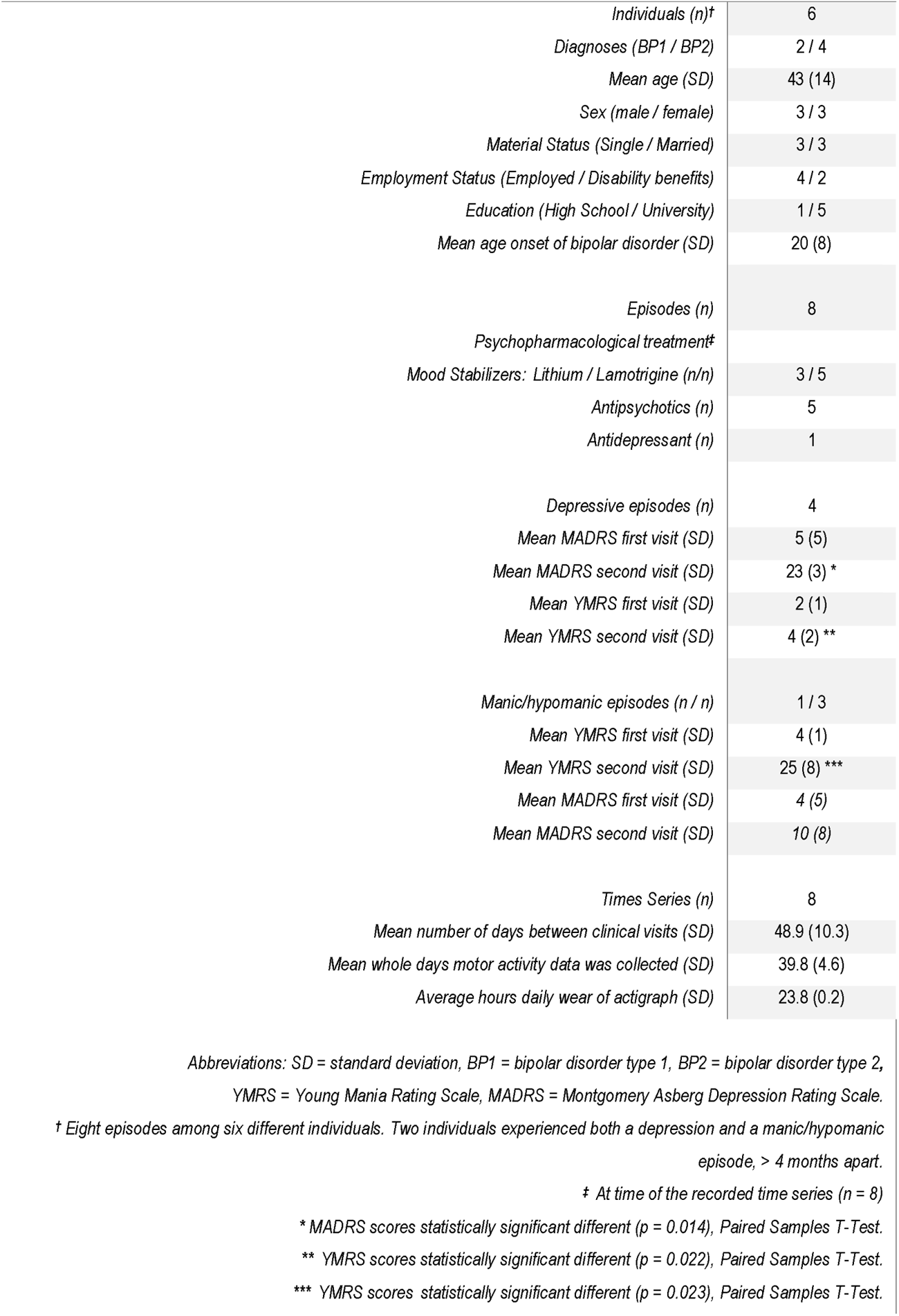
Participant characteristics and demographics, description of mood episodes and motor activity time series.

Forty-nine participants were included in the outpatient study. Thirty-two participants (65 %) provided continuous recordings of motor activity throughout the entire one-year follow-up.

Although 17 of the participants either withdrew (n = 8) or were lost to follow-up (n = 9), 37 participants (76 %) delivered data for more than 6 months. Thirteen participants experienced new mood episodes during follow-up, reporting 19 potential episodes (9 depressive, 9 hypomanic and 1 manic episodes). Of these 19 episodes, one depressive and four hypomanic episodes were undocumented, as these episodes arose and subsided between visits. Four mood episodes were not recorded properly due to extended time gaps between visits, during which the accelerometer battery wore out. These errors occurred partly due to participant non-compliance or were caused by COVID-19-related community lockdowns. Ultimately, ten mood episodes among eight participants fulfilled the criteria of having occurred between two consecutive clinical assessment points not exceeding 2 months (60 days) and still ongoing at the second assessment. However, for one depressive episode, the participant reported not having worn the actigraph, and for one of the hypomanic episodes, the actigraph malfunctioned shortly after the first assessment point, resulting in a lack of available data for two time series. Consequently, eight episodes among six individuals were included in the analysis, four depressions and four (hypo)manic episodes. Two participants are represented with both a (hypo)manic and depressed episode, occurring more than 4 months apart. The participants wore the actigraph on average 23.8 ± 0.2 (mean ± standard derivation) hours each day, for an average of 39.8 ± 4.6 whole days, range 32 to 46 days. Further details about included episodes and participants are given in Table 1.

The output of the LS-USS algorithm, the CACs, and the detected change points are presented in Fig 2. Observing the figure; the algorithm has in most cases segregated the time series into three segments of various statistical properties, divided by the detected change points. Based on the inclusion criteria of euthymic mood state at the start of the motor activity recordings, and in an ongoing mood episode at the end of recordings, we consider the time series segment flanked by the first and the last change point as recognized critical transition periods preceding tipping points. Based on this assumption, the identified critical transitions lasted 13.5 ± 7.2 days (Table 2), depressive episodes lasted 11.4 ± 1.8 days, and (hypo)manic episodes lasted 15.6 ± 10.2 days. However, an additional change point is identified within the alleged critical transitions in three cases (Fig 2: D2, D4, M2). The self-reported time frames of possible mood shifts are marked as Labels in Fig 2 and described in Table 2, which also presents an overview of the detected change points.

**Fig 2.**
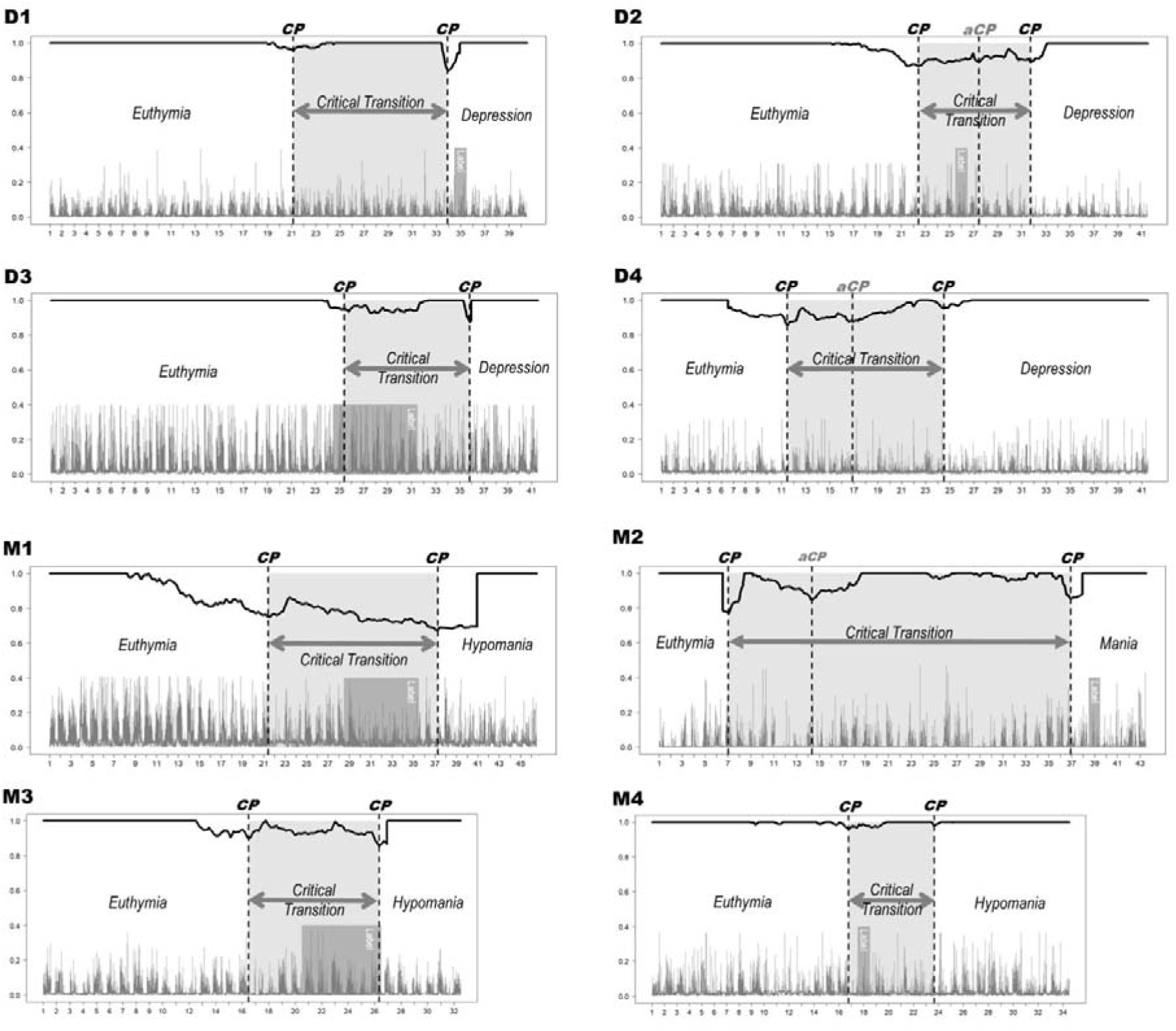
Change points identified in the Corrected Arc Curves produced by the Latent Space Unsupervised Semantic Segmentation algorithm, when analyzing motor activity time series containing mood state changes from euthymic to either depression (D1-D4) or hypomania/mania (M1–M4). *Abbreviations: CP = change points, aCP = additional change points. Label represents the self-reported possible dates/periods for when the change in mood might have occurred. X-axis represents the timeline, with the time marker indicating 12.00 (noon) for each day. Y-axis represents the values of the Corrected Arc Curve (CAC) at the top, and gravitational force equivalents (g) (mean activity) on the bottom, as both variables are expressed as numbers between one and zero. Keep in mind that mean activity is not part of the results, this feature is reported for illustration only*.

**Table 2.** Overview of participant-reported possible occurrences of their mood relapses (labeling), as well as summary of detected Change Points (CP) and the Critical Transitions (CT) observed in the motor activity time series.

|  | Comments<br>participant-reported labeling | Start CT<br>CP<br>(day / HH:MM) | End CT<br>CP<br>(day / HH:MM) | Additional<br>CP<br>(day / HH:MM) | CT<br>Length<br>(days) |
| --- | --- | --- | --- | --- | --- |
| **D1** | Certainly, woke up depressed on day 35. | 21 / 15:30 | 34 / 10:10 |  | 12.8 |
| **D2** | Uncertain, but perhaps depressed from around day 26. | 22 / 21:55 | 32 / 05:25 | 27 / 23:15 | 9.3 |
| **D3** | Uncertain, but depression may have started between day 25 and day 31. | 25 / 21:35 | 36 / 08:00 |  | 10.4 |
| **D4** | Unable to recall. | 11 / 23:45 | 25 / 00:45 | 17 / 10:10 | 13.0 |
| **M1** | Uncertain, but hypomanic episode may have started between days 29 and 35. | 21 / 22:05 | 37 / 18:05 |  | 15.8 |
| **M2** | Very uncertain but was hospitalized at day 39 for a manic episode without psychotic symptoms. | 07 / 13:05 | 37 / 10:00 | 14 / 19:40 | 29.9 |
| **M3** | Unable to recall, but the hypomanic episode may have started between days 21 and 26. | 17 / 00:05 | 26 / 20:20 |  | 9.8 |
| **M4** | Very uncertain, but the hypomanic episode may have started around day 18. | 17 / 06:50 | 24 / 04:00 |  | 6.9 |
| Learning Mean days CT preceding depressions (SD) |  |  |  |  | 11.4 (1.8) |
| Mean days CT preceding hypomania/mania (SD) |  |  |  |  | 15.6 (10.2) |
| Mean days CT preceding all mood episodes (SD) |  |  |  |  | 13.5 (7.2) |

## Discussion

In our study, we observed EWS in motor activity time series preceding bipolar mood state changes, with an average duration of 13.5 days, and a standard derivation of 7.2 days. This finding is in accordance with the objective of this study; to identify critical transition periods preceding bipolar relapses. Preceding depressions, critical transitions are of uniform shorter length (11.4 ± 1.8 days), while those preceding (hypo)manic relapses are less consistent (15.6 ± 10.2 days). Presumably, our findings confirm that the universal theory of complex dynamical systems applies to humans with bipolar disorder experiencing an impending mood episode. Accordingly, our methods and discoveries hold a pronounced potential for future management of bipolar disorder, as the innovative foundation of a new clinical tool. Due to the destructive and potentially lethal consequences of severe bipolar mood episodes,^45^ early intervention and prevention would be beneficial for both patients and clinicians.

However, such a declamatory conclusion may not be fully supported by our findings. In fact, we detected additional change points in three of the time series (Fig 2: D2, D4, M2). Not easily explained, they are cause for speculation. First, the additional change points could be associated with reduced data quality, caused by the 10 Hz recording of motor activity. Normally, motor activity records in higher resolution,^33,46^ as the human activity frequency varies between 0 to 20 Hz, although most human activities happen below 10 Hz. However, according to the Nyquist–Shannon sampling theorem, the sampling rate needs to be twice the frequency of the original signal to obtain full resolution.^29^ Consequently, with 10 Hz recordings, some information is lost, possibly causing a more fragmented stepwise CAC, reporting additional change points. In our case, the 10 Hz sampling frequency was chosen strictly to minimize the battery load and increase maximum recording time, ensuring study feasibility. To our knowledge, no wrist-worn accelerometer with comparable battery capacity able to record at a higher Hz-rate exists on the market. Alternatively, as a second speculation, we could have observed a stepwise development in the critical transition, though similar phenomena are not described in the literature. Still, a stepped development explanation would fit with the observed development within every time series containing additional change points. Finally, the M2 time series could be interpreted as showing unstable behavior from the start. Given that subthreshold symptoms can be observed for over a month prior to an impending mood episode,^3^ a critical transition period could be of corresponding length. In any case, this was not examinable, as the study design did not facilitate additional information at the start of the critical transitions. Regarding the tipping point, or the final change point detected in each time series, most participants were quite uncertain about the timing of the tipping point. Contrastingly, D1 was certain they woke up depressed on day 35, less than a day after the tipping point was indicated. Although M2 had no clue when the tipping point to mania had happened, M2 was hospitalized for mania less than two days after the detected change point (Fig. 2; Table 2). This concurrence between detected change points and the participants’ stories can amplify the LS-USS algorithm’s credibility. Still, none of the other detected tipping points could be verified or falsified. Overall, we evaluate the algorithm to present a presumably sound technique for forecasting approaching shifts in mood, at least when processing off-line batch data.

We analyzed the time series evolving from euthymia to episodic disorder with an unsupervised machine learning algorithm able to identify patterns in undescribed data.^35^ Unsupervised indicates that the algorithm has analyzed the time series blind, and that outcomes are solely based on the evolution of intrinsic static characteristics within each processed time series. The methodological approach of extracting recognized EWS indicators from longitudinal objective real-life data distilled through an autoencoder for dimensionality reduction represents no threat to the integrity of the data.^47^ This contrasts with common handling of multidimensional data within machine learning, where each dimension processes independently before taking the average likelihood over all dimensions at the decision. As some dimensions might not contain helpful information or are heavily correlated with others, the multidimensional average may be diluted. When applying dimensionality reduction by autoencoders, the problem of redundant and correlated information between dimensions diminishes.^35^ Hence, based on the presented hypotheses and evidence that suggest human consciousness can be observed in activity patterns, we propose that EWS could be observed in a CNS-driven pulse wave in the autoencoder-processed one-dimensional signal. This could potentially represent a dissolving euthymic equilibrium switching to another equilibrium – an essential message from the hypothalamus, concealed in the multi-dimensional signal of the EWS indicators.

This study provides valuable insights into the specific use of the LS-USS algorithm. Although a broader perspective is absent, like comparing the algorithms’ efficiency with other similar algorithms. However, other CPD algorithms do not utilize autoencoders, depend on labeled training data, and are consequently ill-suited for unsupervised handling of unlabeled multimodal time series.^34,35^ Similar reservations apply to reservoir computing,^48^ a flexible, supervised, neural network, developed for identifying chaotically transitions and tipping points by reviewing labeled nonlinear complex data. As the underlying objective of this study was to contribute to improving the management of bipolar disorder, we anticipated to operate an unsupervised method capable of forecasting unforeseen future happenings in constant flowing real-time data. An alternative method for consideration could be datamining approaches like anomaly detection algorithms, detecting outliers in the time series.^49^ Such anomalies could be considered EWS, especially if analyzing autoencoder processed EWS indicators. However, as such algorithms are prone to identify only ephemeral changes in the signal, a CPD approach appeared more applicable and competent to identify critical transition periods.^35^

It may seem methodologically unsophisticated and ineffective that, despite applying an advanced state-of-the-art machine learning algorithm, the output of the algorithm is only a single curve, needing manual inspection to identify change points. Reporting and comparing the individual values of the change points are meaningless, as the change point values themselves are just the minimal number in a string of numbers, not statically comparable to the other participants’ change point values.^34^ Normally, accuracy testing is the method for evaluating machine learning calculations.^46^ However, as this method tests the correctness of predictions compared to the empirical evidence, labeled training data are required. Although specific methods for evaluating the accuracy of CPD algorithms have been developed, such methods only apply to results of supervised learning in described data.^34^ Unquestionably, an automated critical transition detection from the CAC procedure is required to enable the innovative potential of this study. This could be done by adjusting the CAC into a standardized scale with a mean of zero and a standard deviation of one. Such normalization is accomplished using a moving window to calculate rolling mean and standard deviation values. Importantly, given the unsupervised nature of the algorithm, the potential change points detected are indicative of any behavioral changes reflected in the actigraphy recordings. Specifically, in bipolar disorder monitoring, critical transition periods are expected to manifest as gradual fluctuations in the CAC, rather than as abrupt or short-lived changes. Accordingly, not all fluctuations below a predefined threshold signify relevant change points. An additional criterion, specifically designed to exclude transient disturbances in the CAC, prioritizing deviations that are sustained and fall below the threshold should be included. This approach is grounded in the understanding that significant behavioral transitions, particularly in bipolar disorder, are gradual and typically extend beyond 12-24 hours. Thus, such an approach to automated critical transition detection from the CAC offers a more precise and meaningful representation of change, recognizing that genuine shifts in behavior unfold over time and are not just short-lived anomalies. This automated approach will be explored in future works.

### Limitations

The most pertinent limitation to our study is that we’ve analyzed a rather small sample of individuals, the sample is also skewed as most participants are diagnosed with bipolar disorder type II. However, there is no prior knowledge indicating that any differences between type I and II should be expected when evaluated as a complex dynamical system. The elevated MADRS scores observed for the (hypo)manic group during the relapse presumably represent dysphoric features, frequently observed in (hypo)manic episodes.^50^ No differences between euphoric or dysphoric (hypo)manic episodes should be expected, as both conditions are characterized by increased energy.^22^ The same group had slightly increased MADRS scores at the start, however, such residual symptoms are commonly observed in euthymic bipolar patients,^51^ and should not destructively affect the representativeness of the sample. Apart from these considerations, we evaluate our study to be of substantial and solid quality, as we analyzed a sample of high clinical accuracy, relevance, and consistency, entirely in line with recommendations for studies on complex dynamical systems in bipolar disorder.^21^ Furthermore, the analyzed datasets are rather complete, averaging 0.8 ± 0.8 percent missing data in the time series, a highly acceptable level of incompleteness.

The small size sample could be relevant for missing observations of tipping points without prior EWS, as observed in the study analyzing daily self-reports of bipolar mood.^18^ However, these missing observations could be related to the contents of the subjective data analyzed. Roughly analogous developments are presented in a simulation study of changeover to depression,^15^ theorizing that abrupt-starting critical transitions moving rapidly towards a tipping point might occur as a response to critical life events. Nevertheless, such happenings would be dependent on the resistance of the system, defined as the flexibility and sturdiness of a current equilibrium to withstand tipping into another equilibrium.^13^ Clearly, a dramatic and critical life event could affect resilience, though this is outside the scope of the current study. Still, one could speculate that the large standard deviation observed, especially before (hypo)manic episodes (Tab. 2), could indicate a variance of resistance in the sample.

## Conclusion

In the present study, we have analyzed motor activity time series of participants with bipolar disorder experiencing mood transitions. Rooted in a complex dynamical systems model, we have applied a scalable, unsupervised domain agnostic change point detection algorithm tailored to handle the specific complexities and nuances of motor activity data in bipolar disorder. The algorithm successfully identified critical transition periods preceding relapses. However, our findings need replication in a larger sample size, as a greater part of the algorithmic detected change points could neither be verified nor falsified, due to lack of time series descriptions. Furthermore, this study lays the groundwork for future research where comparative analyses could be relevant and fruitful, especially as the field evolves and other algorithms are adapted or developed for similar purposes. Going forward, subsequent studies should build on our findings by including comparative analyses to further refine and optimize the early detection of mood state changes.

In conclusion, the LS-USS algorithm seems capable of recognizing impending mood episodes in continuously flowing data streams, at least in pre-collected time series. Accordingly, these results presumably present an innovative method for forecasting approaching mood shifts, facilitating improvement in the clinical management of bipolar disorder.

## Acknowledgments

We acknowledge and thank biostatistician and philosopher Christoffer Andreas Bartz-Johannessen for statistical consultation and for assistance with Fig 2. We would also like to acknowledge research nurse Torkild Hjelmtveit for periodic relief in study participant follow-up. This publication is part of the Introducing Mental Health through Adaptive Technology (INTROMAT) project and was founded by the Norwegian Research Council (agreement 259293).

## Data Availability Statement

The data that support the findings of this study are available on request from the corresponding author. The data are not publicly available due to privacy or ethical restrictions.

